# Facilitators and barriers to the implementation of the WHO ANC-8 contact model among pregnant women: a systematic review in low- and middle-income countries

**DOI:** 10.1101/2025.09.21.25336293

**Authors:** Aschale Worku, Wubegzier Mekonnen, Samson Gebremedhin, Gudina Egata

## Abstract

**Background:** By 2016, the World Health organization (WHO) recommended increasing antenatal care (ANC) contacts from four to a minimum of eight contacts to improve maternal and newborn health outcomes. While barriers to the previously focused ANC model were well documented, facilitators and barriers to this new recommendation are limited. This systematic review synthesized available evidence from low- and middle-income countries (LMICs).

**Methods:** We searched PubMed and Google scholar for studies published between 2016 and 2025 through predefined search terms and Boolean operators. Eligible studies examined facilitators or barriers to the new ANC eight contact model among pregnant women. Quality appraisal was conducted using the JBI-2024 critical appraisal tool individually. Given the heterogeneity, findings were narratively synthesized.

**Result:** seventeen studies met the inclusion criteria, (eight quantitative, nine qualitative). ANC 8+ **c**overage remained low across LMICs, typically between 8-20% with higher uptake in urban over rural. (e.g, Benin 8%, Nigeria urban 35%, Gahana 44%). Barriers included low awareness, financial constraints, limited autonomy in decision making cultural and religious beliefs, Lack of integration of traditional and religious practices into healthcare, poor health system responsiveness and inadequate health insurance coverage. Reported facilitators were higher maternal and partner education, early ANC initiation, male involvement, supportive family, community outreach and media exposures.

**Conclusion:** Uptake of the ANC eight and more contact model remained suboptimal with variations across socio-economic, cultural and health system barriers. Disparities by residence, educational, health insurance highlight the need for equity focused interventions. Strengthening health system, promoting women empowerment and implementing culturally sensitive strategies are critical. Country specific research and fidelity studies are needed to guide effective adaptation of the model in diverse contexts.

## 1. Background

Antenatal care (ANC) is the comprehensive set of health interventions delivered by skilled healthcare professionals to pregnant women and adolescent girls during pregnancy to prevent, detect, manage diseases and promoting healthy behaviors and safe child birth(1,2). Beyond the maternal and newborn care, it also serves an entry point to the broader reproductive health services including health education disease prevention and management of concurrent conditions (2–4).

The origin of ANC trace back to the early 1900’s in Europe(5) which evolved over time to focused antenatal care(FANC) model launched at 2002 (2). World Health organization (WHO) by 2016 recommended pregnant women to receive a minimum of eight ANC contact. In line with it, Ethiopia adapted and started to implement in 2022(3).

Despite the long history of implementing the ANC services globally, the coverage and quality of ANC particularly in low- and middle-income countries (LMICs) is very low. A multi country analysis reported a pooled prevalence of only 13% for ANC eight or more contacts, with a wide variation ranging from 74% in Jordan, 43% in Gahana and Albania (30.0%) to less than 10% in countries such as Senegal, Uganda, and Zambia(6). In Ethiopia, while 74% of pregnant women receive antenatal care first visit, only 43% complete the 4th visit. Furthermore, only 28% of them initiated ANC in the first trimester, despite WHO recommended early ANC initiation before 12 weeks of gestation and complete eight ANC contacts. Alarmingly 32% of pregnant women had their first visit during the fourth or fifth month of pregnancy and 12% had their first visit during the sixth or seventh month) (7) with a low ANC content of care report and facility readiness(8). Population based effective coverage study showed that ANC 4 visit was 40% with regional discrepancies, lower in Afar and higher in A.A. (9). Routine health information report showed that only 27% of pregnant women nationwide receive eight or more ANC contacts(10).

These low coverage and discrepancies indicated a persistent health system constraint alongside the socio-cultural and economic barriers that collectively hinder the uptake of quality ANC services. While many studies have explored the previous FANC model, limited evidence exists about the facilitators and barriers in the adherence of the new ANC eight contact model highlighting that there is insufficient evidence in guiding policy makers and managers for its effective implementations. Therefore, a systematic review is necessary to synthesize existence evidence, identify barriers and facilitators to provide insights for further improving the implementation of the new ANC eight contact model.

## 2. Methods

A scoping review approach was initially undertaken to map the availability of evidence on barriers and facilitators to the new antenatal care (ANC) eight contact model among pregnant women to inform the refinement of the systematic review protocol. The review adhered to the PRISMA 2020 guideline but not registered in Prospero due to the timing constraints.

### 2.1. Search strategy and Eligibility

Data bases searched included PubMed, Google scholar, gray literatures sources-WHO global index, Government reports guided by the PICo framework:

**Population:** Pregnant women

**Phenomenon of interest:** barriers and facilitators to the ANC eight contact model

**Context:** Global

Boolean operators and medical subject headings (MeSH) were applied with a sample PubMed search string attached to (***Appendix 1***).

Studies focusing either on barriers or facilitators to the new ANC eight contact uptake, focused on pregnant women or postnatal women, either qualitative, quantitative studies or mixed studies published after the WHO ANC eight contact model recommendations were included. While studies with primary subjects of policy makers, health care workers, and with wider confidence interval and non-English papers were excluded.

### 2.2. Data extraction and quality assessment

A standardized extraction form was used to capture the Author, year of publication and country, study design and sample size, research aim and context, barriers and facilitators, and key outcomes related to ANC uptake. The methodological quality of studies was assessed using the 2024 JBI guideline-JBI checklist for cross-sectional, and mixed method appraisal tools and its quality assessment was done by an individual.

### 2.3. Data synthesis

Given the expected heterogeneity, a narrative synthesis was employed, and findings were categorized in thematic areas – health system, socio-cultural, individual level factors. Similarities and differences between low- and middle-income countries and high-income countries were highlighted.

### 2.4. Study selection

A total of 110 articles were identified through databases −32 from PubMed and 78 from Google scholar, 4 duplicates were removed, screening 106 titles and abstracts excluded 75 irrelevant studies leaving 33 full texts. Sixteen were excluded (3-inaccessible full texts, 3-pre-dating the WHO ANC eight contact model recommendations, 2-gray literatures, 1-prior FANC model, 1-wider confidence interval and 6-pooled studies from EDHS) leaving the final 17 studies-eight quantitative and nine qualitative met the inclusion and reviewed.

## 3. Results

### 3.1. Characteristics of the studies

A total of 17 studies were included in the review. Geographically, one study was from high income country (Australia), one from upper-middle-income country (Lebanon), two from Asian countries (Pakistan and Myanmar) and the remaining 13 studies were from African countries-Ghana, (02), Mali (1), Nigeria (1), Ethiopia(1), Benin (1), Malawi (02) Rwanda(1), Uganda (02), Zimbabwe (1), and Somali land(1).

Of the included studies 9 were qualitative and 08 were quantitative conducted from 2016-2025 reflecting the period after the world health organization recommended pregnant women to begin ANC early before 12 weeks of gestation and receive at least eight ANC contacts to ensure a positive pregnancy experience and improve maternal and child health (2).

**Figure 1.**
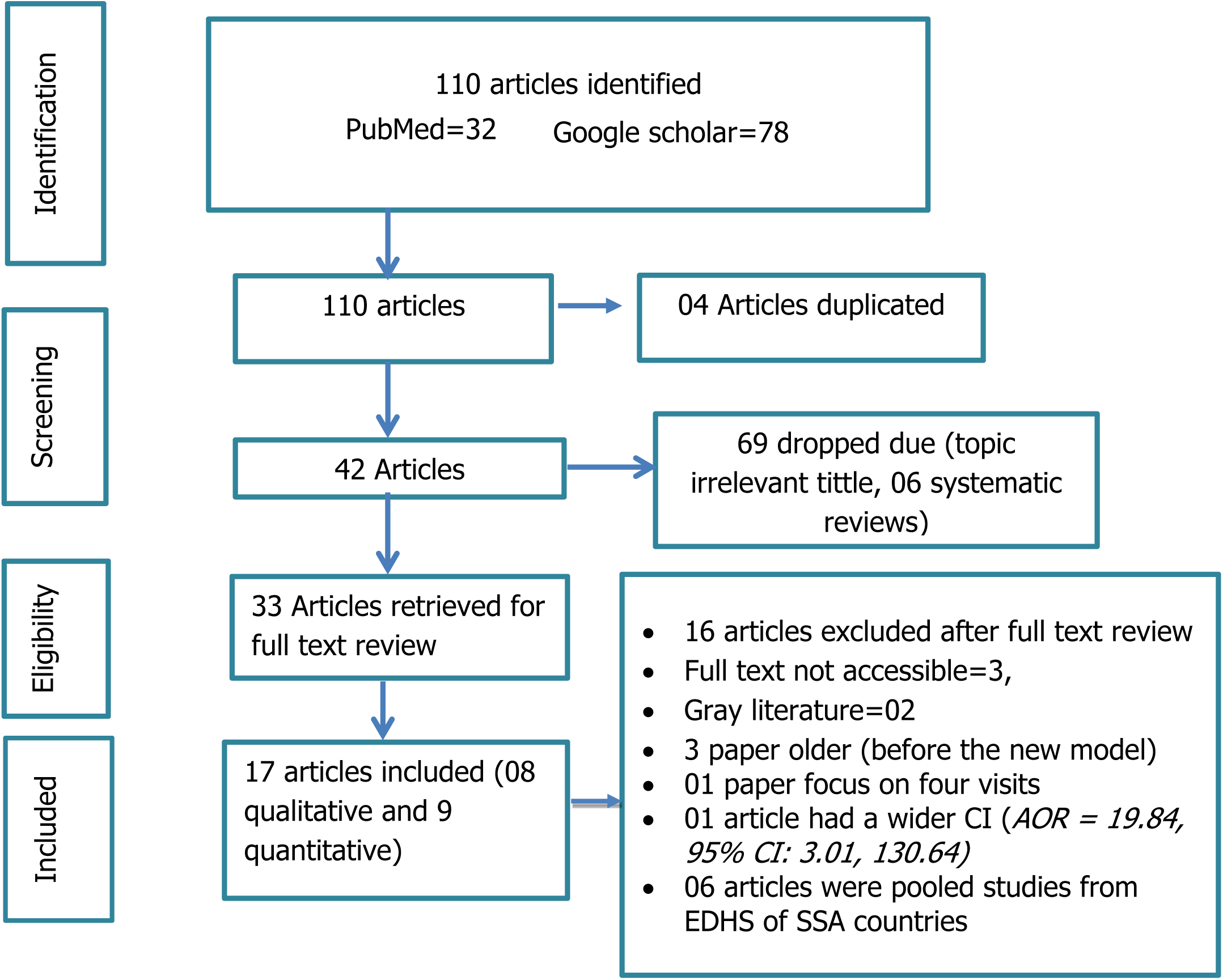
PRISMA-flow diagram for article selection

### 3.2. Quality assessment

Among the 8 quantitative studies 2 studies scored 100%, 5 scored 75% and 1 scored 87.5% quality based on the JBI analytical cross-sectional study critical appraisal checklist. While of the 9 qualitative studies, 3 scored 100%, 3 scored 90% and the remaining 3 scored 70% indicating overall moderate to high methodological quality.

### 3.3. Magnitude of ANC eight contact coverage

The quantitative findings reveal an alarmingly low ANC 8+ coverage particularly in low- and middle-income countries (LMICs) with notable disparities between urban and rural settings. Across most study areas ANC eight or more contact coverage ranged from 8-20% with a consistent higher coverage in the urban settings over rural(11–16). For instance, in Nigeria (2018 DHS), only 20% of pregnant women received a minimum of eight contacts with a stark gap, between urban(35.5%) and rural populations (10.4% rural)(11). Myanmar report a similar ANC eight or more contact coverage at 18%(11,12). While in Ethiopia (Arbaminch urban population) had 41% coverage comparable to urban Nigeria(11,14). Benin showed the much lower coverage at 8%(95%CI 6.5%, 9.7%)(16). While Ghana had the highest coverage at 44.4%(17).

In addition to the frequent number of ANC contacts-eight or more contacts, the WHO’s 2016 recommendations emphasize the importance of early ANC initiation in the first trimester to ensure an optimal ANC uptake. Evidences also indicated that still lower proportion of women had early ANC initiation in many countries-in Myanmar only about 47% of women had an early ANC initiation (12) despite Ghana had a better coverage of 66%.

### 3.4. Factors affecting women to receive the WHO new ANC eight contact model

#### 3.4.1. Socio-demographic characteristics

Several social-demographic factors were found to influence women to adhere to the new ANC contact model. Four quantitative studies reported that maternal age affected both early ANC initiation and completion of the recommended minimum contacts with older women more likely to book early ANC compared to younger(11,12,17,18). Un acceptability study in Uganda found that primigravida women accept the new ANC eight contact model more compared to multigravida women(13). Five studies showed that educational status of women emerged as an important factor affecting women to receive a minimum of eight ANC contact. Women with higher educational level were more likely to early initiate and complete a minimum of eight ANC contact (11,15–18).

Similar studies from Nigeria highlighted that husband’s education and the support they gave improved the uptake of optimal ANC(11,19,20).

The urban rural divide was the most prominent factor in six quantitative studies both in early booking and completing ANC eight or more contacts. Women living in urban settings had a more chance to complete ANC eight contact (11,12,14–16,18).

Early initiation of ANC before or at 12 weeks of gestation is a determinant factor for receiving optimal antenatal care service and completing the ANC eight contacts; two studies indicated that women who early initiate ANC very early were more likely to adhere to the new ANC recommendations(16,18). Knowledge about ANC services was also reported as a factor with studies in Benin and Ethiopia-the better awareness facilitated uptake of the new model (14,16).

Findings from qualitative studies highlighted barriers including limited awareness and misconception about the new antenatal care contract (ANC) model as barriers to adhering to the model. Four of eight qualitative studies reported insufficient knowledge and misunderstanding as major barriers (21–24). Fear of HIV testing both for the women and partner as a barrier was identified in one study-perceived that the more frequent contacts increased the likelihood of being tested for HIV(22).

Finally, a study from Somali land identified short birth spacing as a barrier for adequate ANC care(21). While another study reported that early recognition of pregnancy facilitated timely initiation and optimal uptake(25).

#### 3.4.2. Socio-economic, religious and cultural factors

Socio-economic, religious and cultural factors were also found notably to influence adherence to the ANC 8-contact model. Three quantitative studies reported that religion affected compliance, Christians and Muslims were having higher odds of completing ANC eight or more contacts compared to other religions,though notable differences between the two groups(11,15,17). Some religious beliefs, viewed pregnancy as a natural and God controlled process, discouraging the medical interventions like ultrasound services was reported in one study(21). Moreover lack of integration of traditional and religious knowledge in healthcare was also identified challenge (22). Additionally one study had highlighted ethnic variation in receiving ANC eight or more times (15).

Women’s autonomy in decision making was another key determinant. Four quantitative studies found that greater women’s autonomy in decision making increased the likelihood of completion of ANC eight contacts(8,11,12,18). Household or individual income played an important role, with the wealthier women, the more likely to access and complete optimal ANC services(11–13,15–18). Limited decision making power and domestic responsibilities were also reported as barriers to adhere to the new ANC model in two qualitative studies (21,25).

Cultural barriers were also noted. One study reported that reliance on traditional healers, and fear of the “evil eye” (associated with early pregnancy disclosure) discourage early pregnancy disclosure and initiate ANC services earlier(21).Two studies also reported that feelings of discrimination, lack of privacy and disrespectful care by HCWs were reported as barriers (23,26). Moreover, another identified family violence discourage adherence to the new ANC model (27).

Financial and socio-economic constraints emerged as key barriers in five qualitative studies(21,23,25–27), alongside transportation costs, long distance travelling and fatigue from to frequent ANC contacts (19) in addition to travelling long distances to access health care services(24).

Conversely facilitators included media exposure which was shown in two Nigerian studies (11,15). Four studies identified male involvement, community awareness both of which were highlighted as enablers across four studies (22,24–26).

#### 3.4.3. Health system factors

Health system related factors were also found to affect adherence to the new ANC eight contact model. Three studies reported that women with the existing health problems were more likely to have frequent contacts compared to those attending only for routine pregnancy checks without complication (14,17,20). Health insurance coverage was also an important factor, studies from Nigeria and Ghana had reported that women with health insurance coverage were more likely to adhere to the new model (11,18).

Barriers related to health care access-which delayed women to receive care-were frequently noted especially in rural areas of Nigeria(11) and in Uganda(13). A study from Myanmar found that pregnant women who get services at the private health facility were more likely to complete eight or more ANC contacts (12).

Negative experience within the health system further limited ANC uptake. longer waiting times, unfriendly staff and inflexible scheduling, poor quality health care services (20,21,23,24,27) and also women’s feelings of being discriminated and disrespect by HCWs, lack of privacy were important barriers (22,26). Additionally, poor provider-client communication and lack of cultural sensitivity limit access were also health system barriers reported to receive optimal ANC care while the availability of adequate number of midwifes was described as facilitators.(27). Other facilitators included the presence of community health workers, availability of media campaign, community outreaches activities (25) as well as availability of lady/female health care providers which encouraged them to adhere to ANC contacts(24).

## 4. Discussion

This study revealed that the uptake of WHO recommended ANC eight contacts differs across low- and middle-income countries with both inter and intra-countries variations. A consistent difference was observed between urban and rural setting, where urban women had a more ANC eight contact uptake compared to the rural counterparts. Despite WHO by 2016 recommending an early ANC booking before or at 12 weeks of gestation and completing ANC eight or more contacts, its coverage remains alarmingly low. Reported coverage in majority of the study areas ranged between 8-20% with the lowest coverage in Benin (8%) (16) and higher in Nigeria (20.3%) (15,28). The more higher coverage was reported in Ghana(44.4%)(17) with the weighted prevalence of 41.9%(18). Moreover, beyond the frequent number of ANC contacts early ANC initiation before or at 12 weeks is very critical, but the pregnant women who started ANC before or at 12 weeks was also low. Lower in Myanmar (47%) and Ghana (66%) reflecting inconsistent alignment with the new ANC model. While overall coverage is low, it was still an improvement compared to the sub-Saharan data which was 4.08 % - likely explained by the older DHS data used in those estimates (29).

A range of socio demographic factors were consistently associated with ANC utilization in accordance with the recommendations. Maternal age, Education level of both the women and their partners influence both the early ANC initiation and completion ANC eight or more contacts (11,12,17,18). The urban-rural divide was particularly also a significant factor in affecting the implementation of the new model. In almost all the studies the urban population had a more ANC 8 or more coverage than their counterparts which was in line with the sub-Saharan Africa study findings(29–31), greater accessibility to health facilities in urban areas added with lower transport costs likely explains this disparity so that women in urban settings could have a more chance to frequently contact the health facility as per the recommendations. Qualitative study reports further supports that poor geographic access remains a major barrier in receiving optimal care (11,24,28). Cultural and religious influences also shaped ANC uptake. While no direct prohibition against ANC is observed studies reported that certain religious beliefs hinder the complete use of ANC eight contacts. Christianity and Islam appear to be associated with higher ANC uptake compared to other religious practices(15,17,21). Some religious norms discourage early ANC initiation or medical interventions such as ultrasound investigations. Ethnic variations similarly reported in some studies, further complicating ANC service utilization(11). Addressing these barriers requires multisectoral collaboration with tailored intervention that are culturally sensitive and inclusive.

Financial barriers emerged as a recurrent challenges across most studies in completion of ANC 8+ contacts (21,23,25–27). Direct costs (service fees, transport costs) and indirect costs (time of work) disproportionately affected the rural women. Evidence from the Nigeria and Ghana studies indicated health insurance improved ANC adherence (11,18). These findings highlight the need for financing strategies such as ensuring free maternal health services or expanding insurance coverage, tailored to country specific contexts. In support with this another study reported that insurance coverage and maternal autonomy were an important facilitators for a better adherence of the new ANC recommendations(28)

Women’s knowledge about ANC was also strongly linked in adherence to the new model uptake. Studies consistently demonstrated that health education, media exposure/media campaign and community outreach as public policy facilitated adherence to ANC contacts (13,14,16,22,24,25). This underscores the integration of health education at each point of health care contacts. Moreover, health care workers beyond the frequent ANC contacts and service provisions should also give due attention to women’s awareness about the importance of each ANC services.

Health system factors also played a critical role. Study reports from Nigeria and Ghana highlighted that the availability of health insurance coverage and quality health service provision were the important factors to improve adherence(18,28). Poor quality of healthcare services remains a barrier, particularly in rural areas where infrastructure, staff shortage and delay in health care impede optimal maternal care(23). Interestingly private healthcare facilities in Myanmar demonstrated higher ANC uptake compared to public institutions, suggesting accessibility and perceived service quality directly influence adherence(12). Health system delays, as reported in Uganda, further complicate the implementation of the ANC model(13). Strengthening health system responsiveness, improving service quality and expanding insurance schemes will be central to scaling up ANC 8+ coverage.

It is also important to highlight that ANC 8+ should not be reserved only for women with preexisting illness. While one study noted that women who had preexisting health conditions tend to receive a more ANC eight plus contact compared to those seeking routine antenatal care(14). The WHO framework emphasizes that every pregnant woman is at risk implying that every woman requires at least eight contacts.

Overall, the evidence demonstrates that for the successful implementation of the ANC eight or more contact models, it requires addressing the barriers-financial, educational, cultural, geographic and individual factors. Facilitators such as health insurance schemes, female autonomy in owning resources and decision making, media campaigns, the availability of female health care workers offer a pathway for improvement(21–24,26,27). Ensuring positive experiences through better infrastructure, high quality care and culturally sensitive services will be critical. More importantly these barriers are not left for health care system, rather require relevant stakeholders in addressing barriers such as financial constraints, access to transportation, ensuring women’s autonomy in decision making and gender equity(21,23,25–27). These challenges can be addressed by empowering women through education, economic resources, equitable domestic task-sharing.

In conclusion, this study underscores the complexity of ANC implementation in LMICs. Improving the ANC eight or more contact coverage and quality will require a comprehensive strategy that simultaneously strengthen health systems, empower women, and adapt services to local socio-cultural realities. Targeted interventions in narrowing the gaps in rural and low-income settings are critical in ensuring equitable maternal health care services.

## 5. Study limitations

This review had strengths, it included studies from diverse low- and middle-income country settings, the findings highlight important inter- and intra-country variations that may inform context-specific policy and programmatic decisions. I had included both qualitative and quantitative study designs.

However, it has a limitation. First, the evidence relied on demand from a side perspective, with limited consideration of the supply side barriers and facilitators. Second, the quantitative studies included cross-sectional study designs limiting causal inference. Finally, the quality appraisal was conducted by a single reviewer, which may have introduced assessment bias.

## 6. Conclusion

This review reveals that the implementation of the new ANC eight contact model remains low across low- and middle-income countries with marked disparities by residence, Education, insurance status. Urban, insured women, and educated were more likely to complete the recommended contacts, while the rural and noninsured groups lag. Improving ANC adherence requires a multifaceted approach through strengthening healthcare infrastructure, addressing financial and informational gaps, enhancing service quality, and promoting gender equity and women’s autonomy. Improving the quality ANC 8+ coverage does not rely only on health system factors rather calls for relevant stakeholders to coordinate especially in empowering women.

## 7. Implications for further study

This study showed that barriers affecting the implementation of new ANC eight contact model among pregnant women varies across countries. Some are common, some others are specific to the country which implies a context specific study to identify the facilitators and barriers. Prior to adoption of the new model addressing and identifying facilitators and barriers is very crucial. A qualitative research to addres mainly on some religious and cultural issues that hinder the uptake of both early ANC initiation and complet uptake is important. Furthermore countries like Ethiopia has no national level study about the implementaion status of the ANC eight contact coverage. Existing studies tried to address only isolated components of the new ANC model, rather than evaluating the implementation of the full scope of the new model.

There for implmentation fidelity studies to asses how the current practices align with the WHO ANC framwork will be better to identify more detail gaps to effectively scale up the mplementation

## Abbreviations or Acronyms

ANC: Antenatal care
WHO: World health organization
PRISMA: Preferred Reporting Items for Systematic Reviews and Meta-Analyses.
RH: Reproductive health
A.A: Addis Ababa
PubMED: Publication Medicine
PNC: Postnatal care
HCW: Health care workers
HF: Health facility
N/A: Not applicable
PICo: population, population of interest, context
CI: Confidence interval
EDHS: Ethiopian demographic and health survey
SSA: Sub-Saharan Africa
DHS: Demographic health survey
AOR: Adjusted odds ratio
HIV: Human immune deficiency virus

## Data Availability

All relevant data are within the manuscript and its Supporting Information files

## Acknowledgment

I extend my deepest gratitude to Addis Ababa University, School of Public Health, and the Ethiopian Ministry of Health for granting me the opportunity to pursue my PhD studies. I am sincerely thankful to Project HOPE–Ethiopia for supporting my PhD journey and for its vital role in strengthening the Ministry’s capacity-building efforts.

Myheartfelt appreciation goes to Dr. Samson Gebremedhin (PhD, Associate professor) for his insightful guidance in the preparation of this report and for his instrumental role in facilitating the coordination of the PhD program.

Finally, I am profoundly grateful to my supervisors, Dr. Wubegzier Mekonnen (PhD, Associate Professor) and Dr. Gudina Egata (PhD, Associate Professor), for their unwavering support, mentorship, and invaluable contributions to the success of this research.

## Appendixes

### Appendix 1. Search strategy

This search was done using a combination search term using Boolean operators

1. (“Feasibility Studies”[Mesh] OR “Feasibility Study” OR “Studies, Feasibility” OR “Study, Feasibility” OR OR “ barrier*” OR “ challenge*” OR “ gap” OR “ constraint*” AND ((y_10[Filter]) AND (ffrft[Filter]) AND (clinicalstudy[Filter] OR clinicaltrial[Filter] OR comparativestudy[Filter] OR randomizedcontrolledtrial[Filter]) AND (fft[Filter]) AND (humans[Filter]) AND (english[Filter])) AND ((y_10[Filter]) AND (ffrft[Filter]) AND (clinicalstudy[Filter] OR clinicaltrial[Filter] OR comparativestudy[Filter] OR randomizedcontrolledtrial[Filter]) AND (fft[Filter]) AND (humans[Filter]) AND (english[Filter])))
2. AND (“new ANC model” OR WHO new ANC model OR antenatal care recommendation OR “positive pregnancy experience” OR 2016 ANC model AND ((y_10[Filter]) AND (ffrft[Filter]) AND (clinicalstudy[Filter] OR clinicaltrial[Filter] OR comparativestudy[Filter] OR randomizedcontrolledtrial[Filter]) AND (fft[Filter]) AND (humans[Filter]) AND (english[Filter])) AND ((y_10[Filter]) AND (ffrft[Filter]) AND (clinicalstudy[Filter] OR clinicaltrial[Filter] OR comparativestudy[Filter] OR randomizedcontrolledtrial[Filter]) AND (fft[Filter]) AND (humans[Filter]) AND (english[Filter])))) AND
3. ((“Pregnant People”[Mesh] OR “People, Pregnant” OR “Pregnant Peoples” OR “Pregnant Person” OR “Pregnant Woman” OR “Woman, Pregnant”) OR (“pregnancy”) AND ((y_10[Filter]) AND (ffrft[Filter]) AND (fft[Filter]) AND (humans[Filter]) AND (female[Filter]) AND (english[Filter]) AND (adolescent[Filter] OR alladult[Filter] OR youngadult[Filter] OR adult[Filter]))) Filters: in the last 10 years, Free full text, Full text, English

